# Clinical outcome and outcome prediction of octogenarians with acute basilar artery occlusion and endovascular stroke treatment compared to younger patients

**DOI:** 10.1101/2023.02.15.23285911

**Authors:** Matthias Anthony Mutke, Amanda Zimmermann, Niclas Schmitt, Fatih Seker, Min Chen, Silvia Schönenberger, Markus A. Möhlenbruch, Martin Bendszus, Charlotte S. Weyland, Jessica Jesser

## Abstract

**Background and Aims:** Elderly patients, especially octogenarians, are underrepresented in recently published studies, that showed a benefit of endovascular stroke treatment (EST) for patients with acute basilar artery occlusion (BAO). We aimed to compare the clinical outcome of octogenarians with BAO and EST compared to younger patients and to identify independent outcome predictors.

**Methods:** This is a retrospective, single-center analysis of patients treated for BAO with EST from 01/2013 until 06/2021 in a tertiary stroke center. Octogenarians (≥ 80 years) were compared to younger patients. Study endpoint was the clinical outcome as per modified Rankin Scale (mRS) 90d after stroke onset. The study groups were compared in univariate analysis and a multivariable logistic regression analysis was performed to define independent predictors for clinical outcome.

**Results:** In this study cohort, 74/191 (38.7 %) octogenarians had a higher pre-stroke mRS (Median, IQR: 2, 1 – 3 octogenarians vs. 0, 0 – 1 younger patients, p < 0.001) and a comparable NIHSS before EST (Median, IQR: 21, 8 – 34 vs. 22, 10 – 38 younger, p = 0.712). They showed a comparable mRS 90d after stroke onset (Median, IQR: 5, 2 – 6 younger vs. 5, 3 – 6 octogenarians, p = 0.194), but less often a good clinical outcome (mRS 0-2: n = 27, 23% younger vs. n = 9, 11.7% octogenarians, p = 0.004). The rate of bad clinical outcome was comparable (mRS 5-6, n = 63, 46.7% younger vs. 39, 50.6 % octogenarians, p = 0.194). Baseline NIHSS was a stable independent predictor for clinical outcome in both study groups (e.g. for bad clinical outcome: in octogenarians OR 1.04, CI 100 – 10.85, p = 0.0019, in younger OR 1.061, CI 1.027-1.098, p = 0.005)

**Conclusion:** Octogenarians with acute BAO eligible for EST are less likely to be functionally independent at 90 days after stroke onset, but the rate of death or severe handicap is comparable to younger patients. The admission NIHSS predicts clinical outcome in both age groups.

## Introduction

Endovascular stroke treatment (EST) is an established first-line therapy for acute ischemic stroke and large vessel occlusion in the anterior circulation. There has been persistent doubt about the benefit of EST for basilar artery occlusions (BAO), which account for approximately 10% of all intracranial target vessel occlusions. After earlier studies could not prove the superiority of EST compared to best medical treatment alone for basilar artery occlusions [1, 2], recently presented study results show the contrary[3]. Also, when comparing BAO with anterior circulation large vessel occlusion, the improved outcome after EST is comparable and the intervention is equally safe [4, 5]. As in clinical trials often the case, elderly patients are underrepresented in these studies. However, ischemic stroke has a known age-dependent prevalence with a higher incidence in older age groups [6]. For patients with anterior circulation stroke and EST this gap of knowledge has already been closed, showing that octogenarians and even nonagenarians can still profit from EST [7, 8]. In elderly Chinese patients from the Acute Basilar Artery Occlusion Study (BASILAR), Luo et al. showed, that outcome after EST was worse in patients older than 75 years, but still more effective and safer than conservative treatment [9]. In this study we aimed to determine the clinical outcome of elderly patients presenting with BAO eligible for EST in a western-european stroke center and to define independent predictors for the clinical outcome.

## Methods

The study protocol was approved by the local ethics committee and patient informed consent was waived.

The institutional, prospectively collected, intention-to-treat stroke database was screened for patients who presented with a posterior circulation ischemic stroke due to a basilar artery occlusion (BAO) and eligible for endovascular stroke treatment (EST) between 01/2013 and 06/2021. Stroke etiology was classified according to the TOAST classification: Large-artery atherosclerosis, cardiac embolism, other known causes (e.g. dissection), embolic stroke of unknown source (ESUS) or unknown cause. The extend of final recanalization was rated using the modified TICI scale (mTICI). Additionally, either complete reperfusion of the basilar artery or failed recanalization with remaining occlusion was noted.

### Performance of EST for LVO in the posterior circulation

The decision for EST was made by consensus between neurologist and neurointerventionalist after initial stroke imaging with CT or MRI. Intravenous thrombolysis was administered according to national and international guidelines. The choice of the sedation mode in the complete study cohort was made according to the patient’s compliance, severity of the stroke syndrome and level of consciousness. In the standard approach for EST, a transfemoral access is conducted followed by placing a guide catheter in the subclavian artery (7F / 80 cm Flexor Shuttle, Cook Medical, Bloomington, IN, USA). Subsequently, a distal access catheter is introduced to the vertebral artery (e.g. Sofia 5F, MicroVention, Aliso Viejo, CA, USA). The first line approach (performing contact aspiration or stent-retriever-thrombectomy in combination with continuous distal aspiration using a distal aspiration catheter), as well as the choice of material used for EST was at the discretion of the treating neurointerventionalist. The following stent-retriever types were used (descending order of frequency of use): Solitaire (Medtronic, Dublin, Irland), Trevo (Kalamazoo, Michigan, USA), pRESET (phenox, Bochum, Germany).

### Inclusion and exclusion criteria

All patients eligible for EST of the posterior circulation involving the basilar artery on pre-interventional imaging were assessed for this study. Patients with spontaneous recanalization of the target vessel occlusion after groin puncture (e.g. after i.v. thrombolysis) were excluded. For the outcome analysis patients with missing mRS 90 days after stroke onset were excluded.

### Study endpoints

The primary study endpoint was clinical outcome 90 days after stroke onset measured by the modified Rankin Scale (mRS). As good, favorable and bad clinical outcome according to mRS, we defined 0-2, 0-3, 5-6 respectively. Secondary, a multivariable regression analysis involving clinical, imaging and interventional parameters was performed. Based on the p-values of univariate analysis (p < 0.05) parameters were included in the multivariable regression analysis.

### Statistical analysis

Statistical analysis was performed with GraphPad PRISM (Version 9.0.1). Normality tests were performed, and statistical tests chosen accordingly. Patients were classified into non-octogenarians and octogenarians (80-89 years old) at the time of stroke onset.

Groups were compared for differences in stroke etiology, clinical outcome after 90 days and thrombectomy technique with a Kruskal-Wallis test with Dunn’s correction for multiple comparisons or the Mann-Whitney U test when combining groups and comparing two groups only. Frequency of successful recanalization, type of first thrombectomy attempt and frequency of stenting across different occlusion groups were compared with a Chi-Squared test for multiple groups or Fisher’s exact test for two groups in case groups were combined.

## Results

In this study cohort, octogenarians were more often female compared to younger patients (male sex, n (%), 31 (42%) octogenarians vs. 76 (65%) younger, p = 0.027). They also had a higher posterior-circulation ASPECTS before endovascular stroke treatment (pcASPECTS, median (IQR), 10 (8-10) octogenarians vs. 9 (7-10) younger patients, p = 0.0137). On admission, they had a comparable neurological deficit (NIHSS, median (IQR), 21 (10-38) octogenarians vs. 20 (8-35) younger, p = 0.4867), while they showed a higher pre-stroke disability as per modified Rankin Scale (pre-stroke mRS, median (IQR), 2 (1-3) octogenarians vs. 0 (0-1) younger, p < 0.0001) – see table 1. Octogenarians suffered more often of atrial fibrillation before stroke onset (atrial fibrillation, n (%), 37 (50%) octogenarians vs. 37 (32%) younger, p = 0.0145), while other stroke related comorbidities were comparable between study groups.

**Table 1.** demographic, interventional and imaging parameters of octogenarians and non-octogenarians with acute ischemic stroke and basilar artery occlusion eligible for endovascular stroke treatment.

|  | **Octogenarians**<br>**80-89 years**<br>**(n = 74)** | **Younger patients**<br>**< 80 years**<br>**(n = 117)** | **p-value** |
| --- | --- | --- | --- |
| Age, median (IQR) | 84 (82 – 86) | 70 (59 – 75) | **< 0.0001** |
| Sex, male; n (%) | 31 (42) | 76 (65) | **0.0027** |
| Referred to stroke center from other hospital; n (%) | 35 (47) | 73 (62) | 0.0706 |
| Wake-up stroke; n (%) | 31 (42) | 43 (37) | 0.5426 |
| Posterior-circulation ASPECTS on imaging before EST, median (IQR) | 10 (8-10) | 9 (7-10) | **0.0137** |
| Time from symptom onset to recanalization, min (IQR) | 434 (301-702) | 467 (293-967) | 0.4158 |
| Time from symptom onset to groin puncture, median (IQR) | 331 (209 - 576) | 390 (221 - 817) | 0.2138 |
| i.v. thrombolysis; n (%) | 33 (45) | 51 (44) | > 0.999 |
| NIHSS admission, median (IQR) | 21 (10 – 38) | 20 (8 – 35) | 0.4867 |
| Pre-stroke mRS, median (IQR) | 2 (1-3) | 0 (0-1) | **< 0.0001** |
| **Comorbidities; n (%)** |  |  |  |
| Diabetes mellitus Type 2 | 21 (28) | 21 (18) | 0.1058 |
| Arterial hypertension | 67 (91) | 90 (77) | 0.0656 |
| Coronary artery disease | 28 (38) | 29 (25) | 0.0746 |
| Dyslipidemia | 27 (36) | 33 (28) | 0.3337 |
| Atrial fibrillation | 37 (50) | 37 (32) | **0.0145** |
| **Interventional details during EST** |  |  |  |
| Successful recanalization of basilar artery occlusion; n (%) | 64 (86) | 107 (91) | 0.3338 |
| Number of thrombectomy attempts; median (IQR) | 1 (1-2) | 1 (1-3) | 0.6426 |
| Intracranial stenting during EST | 16 (22) | 24 (21) | 0.8571 |
| Stent-retriever thrombectomy as first approach; n (%) | 54 (73) | 79 (68) | 0.3162 |
| Contact aspiration as first approach; n (%) | 10 (14) | 28 (24) | 0.089 |
| Direct intracranial stenting, no thrombectomy attempt; n (%) | 4 (5) | 5 (4) | 0.970 |
| Occlusion not reached; n (%) | 5 (7) | 3 (3) | 0.257 |
| **Stroke etiology; n (%)** |  |  |  |
| atherosclerosis | 23 (31) | 37 (32) | 0.4218 |
| Cardio-embolic | 31 (42) | 36 (31) | 0.127 |
| Other causes | 1 (1) | 4 (3) | 0.393 |
| Embolic stroke of unknown source | 10 (14) | 25 (21) | 0.189 |
| Unknown (work-up not completed) | 9 (12) | 15 (13) | 0.928 |
| **Clinical outcome 90 days after stroke onset per modified Rankin Scale (mRS), n (%)**<br>**Missing values: octogenarians = 6, non-octogenarians = 11** |  |  |  |
| Good outcome (mRS 0-2), | 8 | 17 | 0.485 |
| Favorable outcome (mRS 0-3) | 17 | 30 | 0.725 |
| Bad outcome (mRS 5-6) | 34 | 55 | 0.971 |

ASPECTS = Alberta Stroke Program Early CT Score, EST = Endovascular Stroke Treatment, IQR = inter quartile range, mRS = modified Rankin Scale, NIHSS = National Institutes of Health Stroke Scale, i.v. = intra-venous

Both study groups had the same rate of successful basilar artery recanalization, a comparable rate of intracranial stenting during EST and the first line approach when comparing contact aspiration and stent-retriever thrombectomy did not differ – see also table 1. For octogenarians the leading cause of stroke was cardioembolic with a significantly higher rate of cardioembolic stroke compared to younger patients (n (%), 31 (42%) octogenarians vs. 36 (31%) younger patients). Younger patients showed more often an embolic stroke of unknown source (ESUS) than octogenarians, while atherosclerosis was as likely as cardioembolic stroke as stroke etiology in younger patients – see table 1.

When comparing the outcome, younger patients were more often functionally independent 90 days after stroke onset (mRS 0-2, n (%), 9 (12.1%) octogenarians vs. 27 (23.1%) younger patients) and showed more often a favorable clinical outcome (mRS 0-3, n (%), 18 (24.3%) octogenarians vs. 37 (31.6%) younger patients). For bad outcome, comprising severe handicap or death 90 days after stroke onset, the two study groups showed no difference (mRS 5-6, n (%), 39 (52.7%) octogenarians vs. 60 (51,3%) younger patients) – see also figure 1.

**Figure 1.**
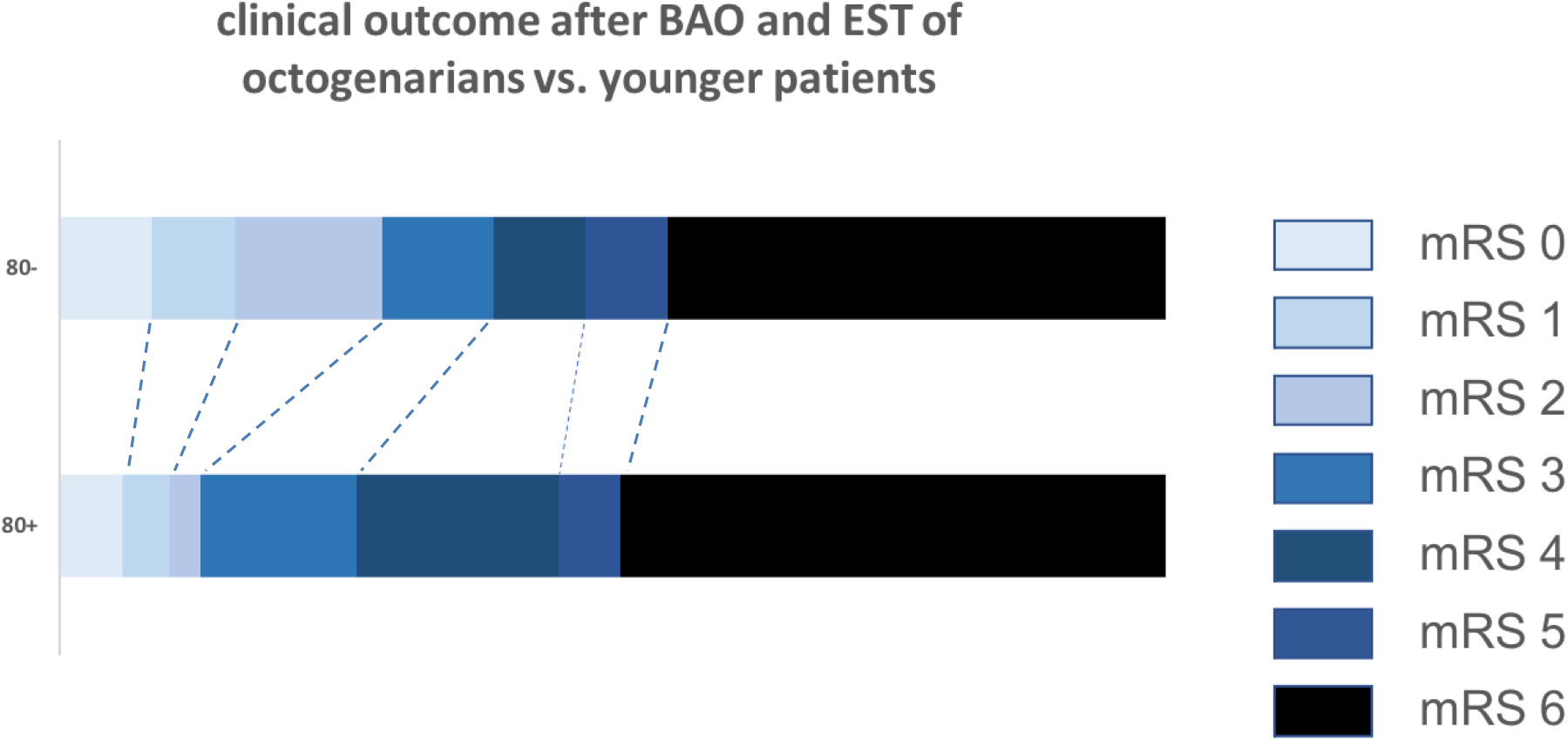
**group comparison of clinical onset as per modified Rankin Scale 90 days after stroke onset of young patients (80-) vs. octogenarians (80+), BAO = Basilar Artery Occlusion, EST = Endovascular Stroke Treatment**

Independent predictors for octogenarians regarding favorable outcome (mRS 0-3 90 days after stroke onset) were the NIHSS on admission (OR 0.919, Confidence Interval (CI) 0.86-0.97, p = 0.0019) and pre-stroke mRS (OR 0.542, CI 0.30 -0.90, p = 0.0291). For bad outcome (mRS 5-6 after 90 days), the NIHSS on admission stayed independently predictive (OR 1.04, CI 1.00-1.085, p = 0.0591) – see table 2.

**Table 2.**
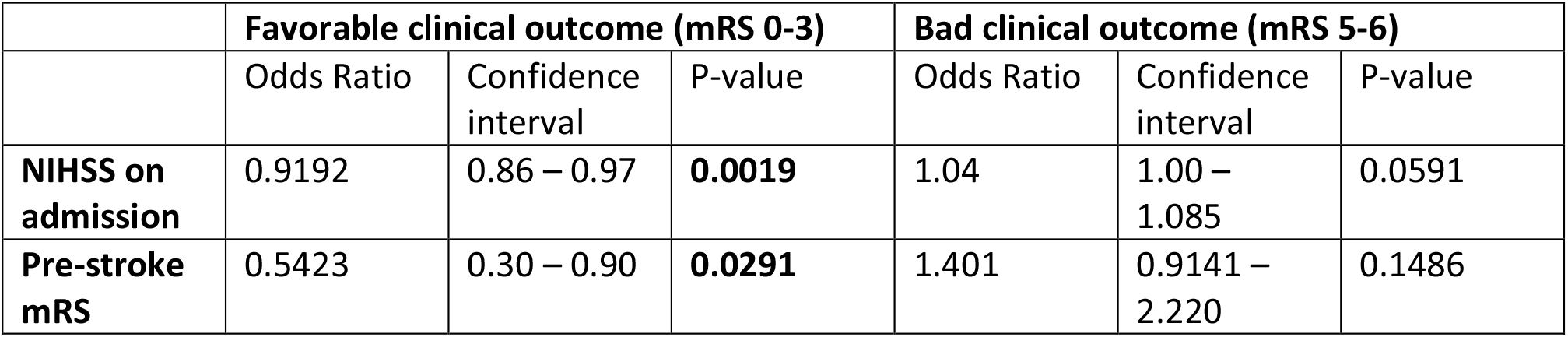
independent predictors of favorable clinical outcome (mRS 0-3) and bad clinical outcome (mRS 5-6) in logistic regression analysis for octogenarians.

|  | Favorable clinical outcome (mRS 0-3) |  |  | Bad clinical outcome (mRS 5-6) |  |  |
| --- | --- | --- | --- | --- | --- | --- |
|  | Odds Ratio | Confidence interval | P-value | Odds Ratio | Confidence interval | P-value |
| **NIHSS on admission** | 0.9192 | 0.86 – 0.97 | **0.0019** | 1.04 | 1.00 – 1.085 | 0.0591 |
| **Pre-stroke mRS** | 0.5423 | 0.30 – 0.90 | **0.0291** | 1.401 | 0.9141 – 2.220 | 0.1486 |

For younger patients, the NIHSS on admission was likewise an independent predictor for favorable outcome (OR 0.935, CI 0.90 – 0.97, p = 0.0002) and bad outcome (OR 1.061, CI 1.027 – 1.098, p = 0.005) – see table 3.

**Table 3.** independent predictors of favorable clinical outcome (mRS 0-3) and bad clinical outcome (mRS 5-6) in logistic regression analysis for non-octogenarians.

|  | Favorable clinical outcome (mRS 0-3) |  |  | Bad clinical outcome (mRS 5-6) |  |  |
| --- | --- | --- | --- | --- | --- | --- |
|  | Odds Ratio | Confidence interval | P-value | Odds Ratio | Confidence interval | P-value |
| **NIHSS on admission** | 0.935 | 0.90 – 0.97 | **0.0002** | 1.061 | 1.027 – 1.098 | **0.005** |

## Discussion

In this study of patients eligible for endovascular stroke treatment with basilar artery occlusion and acute ischemic stroke, octogenarians were less likely to be functionally independent 90 days after stroke treatment and less likely to show a favorable outcome. However, they had the same risk to suffer a very bad outcome (mRS 5-6) compared to younger patients.

These results are comparable to studies referring to anterior circulation large vessel occlusions, that showed that very old patients still possibly profit from EST [10, 11]. Regarding western-european study populations, this is the first study in the era of modern EST investigating clinical outcome for octogenarians after BAO and EST compared to younger patients. Vergouwen et al. showed in 2012 for patients older than 75 years old, that elderly patients can still profit from EST, although the mortality rate was higher for the elderly, which contradicts our study results [12]. This study is comparable to a Chinese population-based study (BASILAR) by Luo et al., which found that patients older than 75 years at stroke onset still profit from EST compared to best medical treatment [9]. In the intervention arm of the BASILAR study, the admission NIHSS was also an independent predictor for clinical outcome, whereas pre-stroke mRS was not investigated in that study, which proved to be predictive for favorable outcome in our study. Contradictive to our study results, there was a higher mortality rate for older patients and the rate of favorable outcome was comparable between age groups. Also contradictive to Luo et al., there were more female stroke patients among octogenarians in our study cohort. This might be due to the overall lower life expectancy (75.6 years in men and 81.3 years in women) in China compared to Germany (78.5 years in men and 83.4 years in women). Further studies concerning sex differences in ischemic stroke treatment and outcome are warranted to define important sex-related differences of this disease. However, Deb-Chatterji et al. found, that sex alone is not an independent predictor for ischemic stroke outcome, but that the worse clinical outcome for women after acute stroke care is crucially confounded by the higher representation of women in the elderly group and the age-associated higher pre-stroke functional status [13]. Luo et al. also found a higher rate of cardioembolic stroke in the elderly, which is in line with our study results. While atherosclerotic disease is overall more prevalent in posterior circulation stroke compared to the anterior circulation, the higher incidence of atrial fibrillation in older patients, especially in women, leads to a higher prevalence of cardioembolic stroke origin [14, 15]. They did not report on the rate of embolic stroke of undetermined source (ESUS) as stroke etiology, which proved to be higher in younger patients in our study cohort compared to octogenarians. ESUS has a known age-related distribution in stroke patients with a higher likelihood in younger patients [16].

Limitations of this study concern the single-center retrospective study design. Especially the distribution of stroke etiology can differ depending on the geographical background of the study population. A population’s life expectancy also influences study results concerning elderly patients and acute ischemic stroke care. Facing the demographic change in western populations, further research is warranted addressing older populations.

## Conclusion

Octogenarians are less likely to achieve functional independency or favorable clinical outcome after basilar artery occlusion and endovascular stroke treatment compared to younger patients. However, the rate of severe handicap or death is alike. The admission NIHSS is an independent predictor for favorable outcome and bad clinical outcome in both age groups.

## Data Availability

All data produced in the present study are available upon reasonable request to the authors

## References

1. Liu, X., et al., Endovascular treatment versus standard medical treatment for vertebrobasilar artery occlusion (BEST): an open-label, randomised controlled trial. Lancet Neurol, 2020. 19(2): p. 115–122.

2. Schonewille, W.J., et al., Treatment and outcomes of acute basilar artery occlusion in the Basilar Artery International Cooperation Study (BASICS): a prospective registry study. Lancet Neurol, 2009. 8(8): p. 724–30.

3. Malik, A., et al., Mechanical thrombectomy in acute basilar artery stroke: a systematic review and Meta-analysis of randomized controlled trials. BMC Neurol, 2022. 22(1): p. 415.

4. Sommer, P., et al., Thrombectomy in basilar artery occlusion. Int J Stroke, 2022: p. 17474930211069859.

5. Meinel, T.R., et al., Mechanical thrombectomy for basilar artery occlusion: efficacy, outcomes, and futile recanalization in comparison with the anterior circulation. J Neurointerv Surg, 2019.

6. Hollander, M., et al., Incidence, risk, and case fatality of first ever stroke in the elderly population. The Rotterdam Study. J Neurol Neurosurg Psychiatry, 2003. 74(3): p. 317–21.

7. Drouard-de Rousiers, E., et al., Impact of Reperfusion for Nonagenarians Treated by Mechanical Thrombectomy: Insights From the ETIS Registry. Stroke, 2019. 50(11): p. 3164–3169.

8. Mohlenbruch, M., et al., Endovascular Stroke Treatment of Nonagenarians. AJNR Am J Neuroradiol, 2017. 38(2): p. 299–303.

9. Luo, W., et al., Endovascular intervention for basilar artery occlusion in the elderly. Ther Adv Neurol Disord, 2021. 14: p. 17562864211000453.

10. Broussalis, E., et al., Endovascular mechanical recanalization of acute ischaemic stroke in octogenarians. Eur Radiol, 2016. 26(6): p. 1742–50.

11. Hendrix, P., et al., Outcome Following Mechanical Thrombectomy for Anterior Circulation Large Vessel Occlusion Stroke in the Elderly. Clin Neuroradiol, 2021.

12. Vergouwen, M.D., et al., Outcomes of basilar artery occlusion in patients aged 75 years or older in the Basilar Artery International Cooperation Study. J Neurol, 2012. 259(11): p. 2341–6.

13. Deb-Chatterji, M., et al., Sex Differences in Outcome After Thrombectomy for Acute Ischemic Stroke are Explained by Confounding Factors. Clin Neuroradiol, 2021. 31(4): p. 1101–1109.

14. Marini, C., et al., Contribution of atrial fibrillation to incidence and outcome of ischemic stroke: results from a population-based study. Stroke, 2005. 36(6): p. 1115–9.

15. Russo, T., G. Felzani, and C. Marini, Stroke in the very old: a systematic review of studies on incidence, outcome, and resource use. J Aging Res, 2011. 2011: p. 108785.

16. Hart, R.G., et al., Embolic Stroke of Undetermined Source: A Systematic Review and Clinical Update. Stroke, 2017. 48(4): p. 867–872.

